# Associations of salivary cortisol with cardiovascular parameters and mortality in the MRC National Survey of Health and Development (NSHD)

**DOI:** 10.1101/2023.11.29.23299189

**Authors:** Lamia Al Saikhan, Dylan M Williams, Gaby Captur, Alun D Hughes, Daniel Davis, Nish Chaturvedi

## Abstract

**Objectives:** Hypothalamic-pituitary-adrenal (HPA) dysregulation is a postulated risk factor for cardiovascular disease but results from previous studies have been mixed. We examined cross-sectional associations of diurnal patterns of salivary cortisol with 10 subclinical measures of cardiovascular disease (including pulse wave velocity, carotid intima-media thickness, and measures of heart structure and function), and prospective associations of cortisol variation with cardiovascular (CV) and non-cardiovascular (NCV) mortality.

**Methods:** We included up to 1,263 participants of the MRC National Survey for Health and Development birth cohort, who had participated in serial diurnal salivary cortisol sampling in 2006-2010 (aged 60–64 years). We used multivariable linear and multinomial logistic regression to estimate associations of cortisol awakening response (CAR), slope and area under the curve (AUC) with subclinical cardiovascular measures, and multivariable Cox regression to estimate associations of the three cortisol metrics with all-cause, CV and NCV mortality.

**Results:** All associations with subclinical cardiovascular measures were weak and most were compatible with the null hypothesis. Each standard deviation (SD) higher cortisol AUC was associated with a 1.9% (95%-CI 0.3-3.5) higher pulse wave velocity. A shallow cortisol slope (per SD gradient) was associated with a 1.4% lower intima media thickness (95%-CI −2.6, − 0.2), and lower odds of concentric remodelling (OR 0.83, 95%-CI 0.70-0.99). Given the multiple analyses performed, these could be chance findings. AUC, cortisol slope and CAR were not convincingly associated with cardiovascular or total mortality, although the 95% confidence intervals of hazard ratio estimates were wide.

**Conclusions:** This study provides little evidence for clinically important associations of salivary cortisol and subclinical cardiovascular measures and mortality.

## 1. Introduction

The hypothalamic-pituitary-adrenal (HPA) axis controls the secretion of cortisol in response to acute stressors and diurnal patterning. HPA axis dysregulation is usually characterised by a blunted cortisol awakening response (CAR, the rapid increase of cortisol after waking),^1–3^ higher overall cortisol secretion and a shallower decline throughout the day.^1,4^ Based on animal and some human research, it has been suggested that repeated and prolonged exposure to stressors across the lifetime is likely to contribute to HPA dysregulation,^5^ implicating the HPA axis in the pathway between psychosocial stress and health.^6,7^

Several studies have suggested a link between diurnal salivary cortisol and cardiovascular mortality, but results have been mixed in terms of which measures of diurnal rhythm showed associations and might have been influenced by limited number of events or short follow-up. Studying subclinical measures as surrogates for clinical endpoints can improve statistical power and reduce selection bias, and may help to elucidate mechanistic actions of exposures. However, available evidence on associations of salivary cortisol excursions and subclinical cardiovascular measures to date is inconclusive and has been limited to measures of atherosclerosis.^10–15^

In this study we aimed to: (1) investigate the cross-sectional associations of diurnal patterns of salivary cortisol with multiple subclinical measures of cardiovascular disease; (2) investigate the prospective associations of diurnal salivary cortisol patterns with cardiovascular and non-cardiovascular mortality in participants of the MRC National Survey for Health and Development study (NSHD), aged 60-64 years at cortisol sampling. We hypothesised that HPA axis dysregulation would be associated with evidence of subclinical cardiovascular disease and cardiovascular mortality.

## 2. Materials and methods

### 2.1. Study Population

We analysed data from participants from the NSHD, a birth cohort of 5362 individuals born Britain in one week of March 1946. At age 60-64 years (2006-2010), participants were invited to a clinic assessment that included health, lifestyle and socio-demographic questionnaires, cardiovascular measures and a serial salivary cortisol collection protocol. Ethical approval for the study was obtained from various regional ethics committees, and participants provided written informed consent for each component of data collection.^16^

## 2. Assessment of diurnal salivary cortisol

Participants provided four saliva samples over 24 hours: i) during their clinic appointment, ii) the same day between 21:00 – 21:30, iii) on waking the following day, and iv) 30 minutes later. At the clinic, a research nurse explained the protocol for home cortisol collection, including requests not to eat, drink or smoke for 30 minutes before each sample collection, and to report any protocol deviations.^17^ Salivettes were returned via post and frozen before radioimmunoassay in Dresden, Germany.^3^ Cortisol collection was not conducted for a subset of participants attending a clinic in Manchester (n=348) because these were part of a previous feasibility sample that did not include cortisol sampling.

We used the four data cortisol points to calculate three diurnal cortisol measures: (1) cortisol awakening response (CAR), which is regulated separately from the diurnal cycle.^18^ This was measured as the difference between the waking cortisol and the sample 30 minutes later; (2) Diurnal cortisol slope, derived from a random-effects linear regression of the log-transformed cortisol across the first, third and fourth cortisol measurements resulting in a negative slope variable with steeper slopes represented by more negative values;^6^ (3) Area under the curve (AUC), using mean time between measurements, to quantify overall cortisol exposure.^2,19^

### 2.3. Outcome assessment

Subclinical cardiovascular measures included carotid-femoral pulse wave velocity (PWV), common carotid artery intima-media thickness (cIMT), and measures of cardiac structure (left ventricular (LV) mass indexed for height,^2.7^ left atrial (LA) volume indexed for body surface area (BSA), LV relative wall thickness (RWT), and LV remodelling categories (Lang et al. 2015), and function (ejection fraction (EF), peak early systolic velocity of the mitral annulus (s’), peak early diastolic mitral annulus velocity (e’), ratio of peak early diastolic mitral inflow velocity to peak early diastolic mitral annulus velocity (E/e’), N-terminal-pro B-type natriuretic peptide (NT-proBNP), a global measure of LV function and independent biomarker for heart failure prognosis.^20^ Cardiac structure and function were measured with echocardiography using a GE Vivid-I ultrasound scanner. Images were recorded from parasternal long axis and short axis views, apical 5-chamber, 4-chamber, 3-chamber, 2-chamber and aortic views (as well as a tissue Doppler in 4-chamber view).^16^ The following cardiac structure parameters were calculated according to recommendations by the American Society of Echocardiography: LV mass indexed for height,^2.7^ relative wall thickness, remodelling categories (normal defined as RWT ≤0.42 and LV mass indexed for BSA of ≤95gm/m2 for women and ≤115 for men, eccentric hypertrophy defined as RWT ≤0.42 and LV mass indexed for BSA >95 for women and >115 for men, concentric remodelling defined as RWT >0.42 and LV mass indexed for BSA of ≤95 for women and ≤115 for men, and concentric hypertrophy defined as RWT >0.42 and LV mass indexed for BSA of >95 or women and >115 for men),^21^ the systolic function parameters EF, s’, and the diastolic function parameters LA volume indexed to body surface area and E/e’. NT-proBNP was measured using automated electrochemiluminescence immunoassay (R&D Systems Abingdon, UK).^23^ PWV was measured using a Vicorder device.^24^ PWV above 30m/s were capped. cIMT was measured bilaterally using a GE Vivid-I ultrasound scanner and calculated using automated software.^25^

Mortality information was available through linked death records until December 31, 2021, a mean 12 years from cortisol data collection. Cardiovascular death was defined as the primary cause coded by International Classification of Disease 10 as I100I199 and 9 401– 454; non-cardiovascular death was defined as any other cause of death. Deaths from unknown cause were excluded from non-cardiovascular death.

### 2.4. Confounders

Noise variables and potential confounders were selected based on recommendations for salivary cortisol analyses and expected associations with cardiovascular outcomes, respectively: waking time, having eaten or drunk before cortisol measurement, leisure time physical activity and education.^2^ We also investigated potential mediation by BMI and overall cardiovascular risk. BMI was measured by trained staff using a standardised protocol; education and leisure time physical activity were self-reported. Cardiovascular risk was quantified by calculation of a Framingham risk score using age, sex, measured HDL and LDL cholesterol, systolic and diastolic blood pressure, diabetes (based on self-report and/or use of antidiabetic drugs or insulin) and smoking).^26^

## 3. Statistical analysis

Participants were excluded from analyses if they had not participated in the saliva sample collection at all four time points, took steroid medications, smoked within 30 minutes of cortisol measurement, missed recording the measurement time, were probably shift-working (waking before 4.00 or after 12.00), took the evening measurement before 20:00, collected samples in the wrong order and if the 30-minute post-waking measurement was done >1h after waking.^17^ Continuous outcome measures were log-transformed to account for skewed distributions in most measures. We undertook complete-case analyses for those with cortisol data and confounder data (variables included in model 1-3, below) since missingness among confounders was less than 6%.

We used multivariable linear regression to estimate the association of CAR, slope and AUC with each cardiovascular structural and functional measure (except where remodelling category was the outcome; here, we used multinomial logistic regression) and Cox regression to estimate hazard ratios (HR) for all-cause mortality, cardiovascular and non-cardiovascular mortality. CAR, slope and AUC were entered in models as continuous variables with units representing per standard deviation (SD) increase. We tested for non-linearity between exposures and outcomes using fractional polynomial models. We multiplied β-coefficients by 100 to yield percentage differences in outcomes per SDs of exposure.^27^ We used four sequential models to assess the effects on estimates of adjusting for potential confounders and noise variables (models 1 and 2) and covariates that could lie on the pathway between exposure and outcome (models 3 and 4). Model 1 was adjusted for sex, waking time (per hour later), eating or drinking before cortisol measurement (yes/no). Model 2 additionally adjusted for education (no qualification/ vocational only/ O level or equivalent/ A level or equivalent/ higher education) and leisure time physical activity (none/ 1-4 times per week/ ≥5 times per week); Model 3 was additionally adjusted for BMI and Model 4 for Framingham risk score.

Based on earlier findings reporting sex differences in the association of cortisol slope and PWV,^14^ we compared models with and without an interaction term by sex using likelihood-ratio tests. We conducted three sensitivity analyses: (1) excluding log cortisol and cardiovascular parameters beyond ±3 SD of mean values to identify the impact of outliers; (2) excluding participants with self-reported clinical diagnoses of myocardial infarction and angina since 1999 (cases n=70) to rule out any role for overt coronary heart disease; (3) excluding any deaths within the two years of follow-up which may mitigate bias from reverse causality.

We did not formally appraise results with a threshold of statistical significance adjusted for a burden of multiple testing, but interpreted any unclear associations cautiously in light of the many exposure-outcome associations tested in these analyses.

## 4. Results

1,669 participants provided saliva samples at all four time points, of which 330 had to be excluded and an additional 78 had missing confounder data – leaving an analytical sample of 1,263 (Table 1). Based on outcome availability, sample sizes ranged from 867 for EF to 1169 for NT-proBNP. There was little difference in heart and vascular measures when comparing included participants to ineligible participants from the wider cohort (Table 1), but those excluded had lower education levels, were more likely to be men and had higher Framingham risk scores (Supplementary Table S1).

**Table 1:**
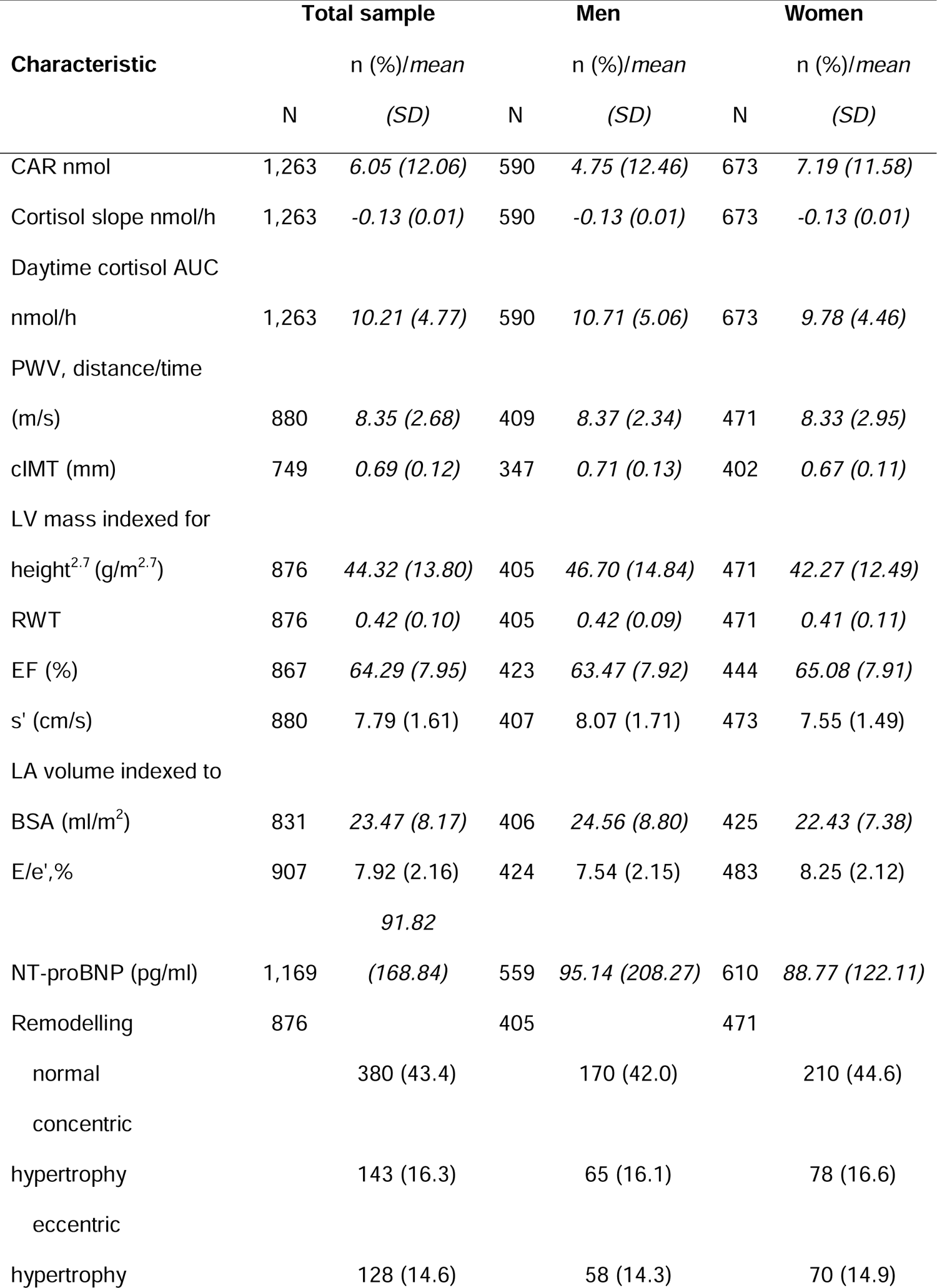
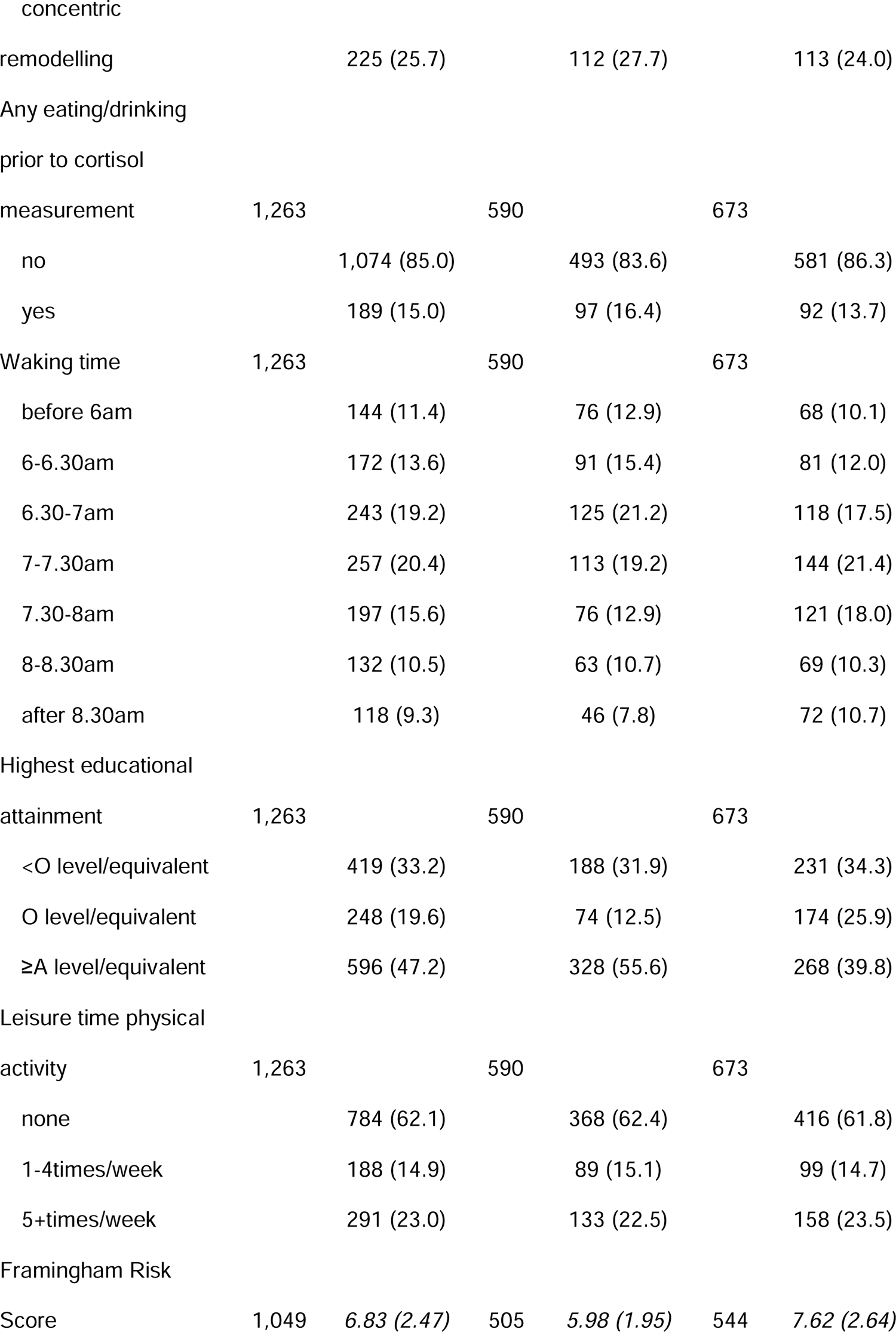
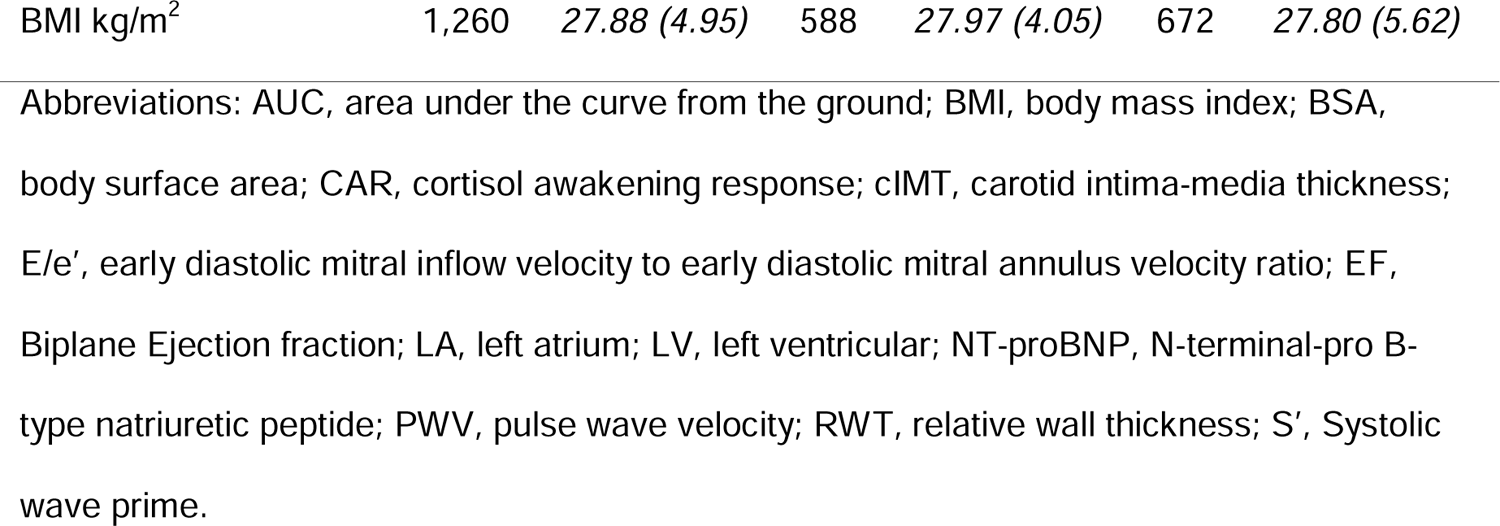
Characteristics for total sample and by sex.

There was no convincing association between cortisol awakening response and any cardiovascular parameter (Table 2). A shallow (i.e. less negative) cortisol slope was associated with a 1.4% lower cIMT (95%-CI −2.6, −0.2; Table 2), and a lower odds of concentric remodelling (OR 0.83, 95%-CI 0.70, 0.99; Table 3) in models adjusted for sex, any eating or drinking, waking time, education, and physical activity. Each standard deviation higher cortisol AUC was associated with a 1.9% (95%-CI 0.3, 3.5) higher pulse wave velocity and a 1.5% (95%-CI 0.0, 2.9) higher s’. These associations did not differ substantially when fewer covariates were included (supplementary Table S2) and mostly remained when additionally adjusted for BMI and Framingham risk scores. The association of AUC and s’ was attenuated with additional adjustment for Framingham risk scores. There was no association between cortisol slope and AUC with any other vascular and cardiac parameter. Associations showed no evidence for non-linearity (*P* values >0.07) or modification by sex (*P* values for interaction >0.05), except CAR and EF (*P* value for sex interaction=0.03, adjusted % difference in men −1.2, 95%-CI −2.5, 0.2; in women 0.8, 95%-CI −0.5, 2.1).

**Table 2:**
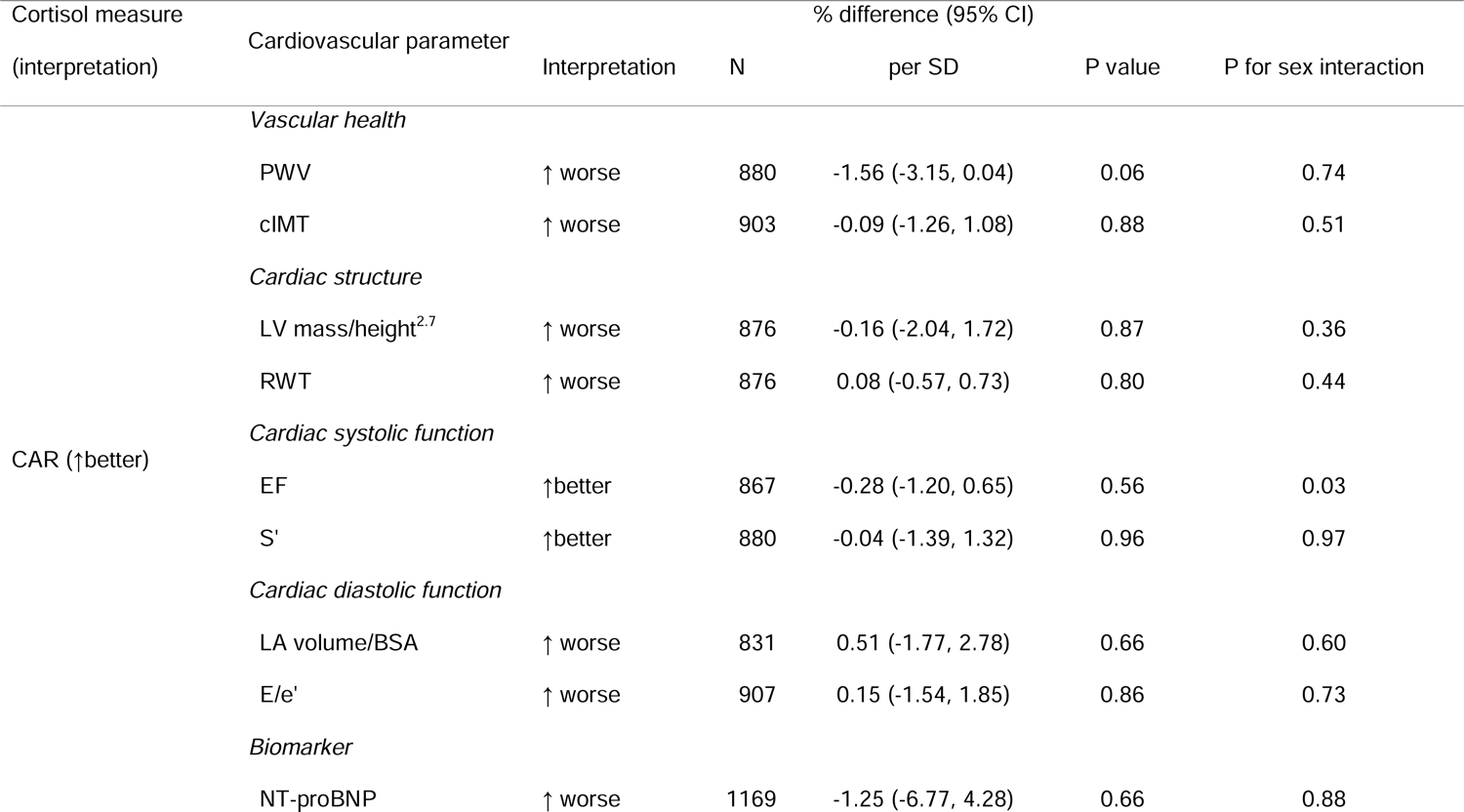
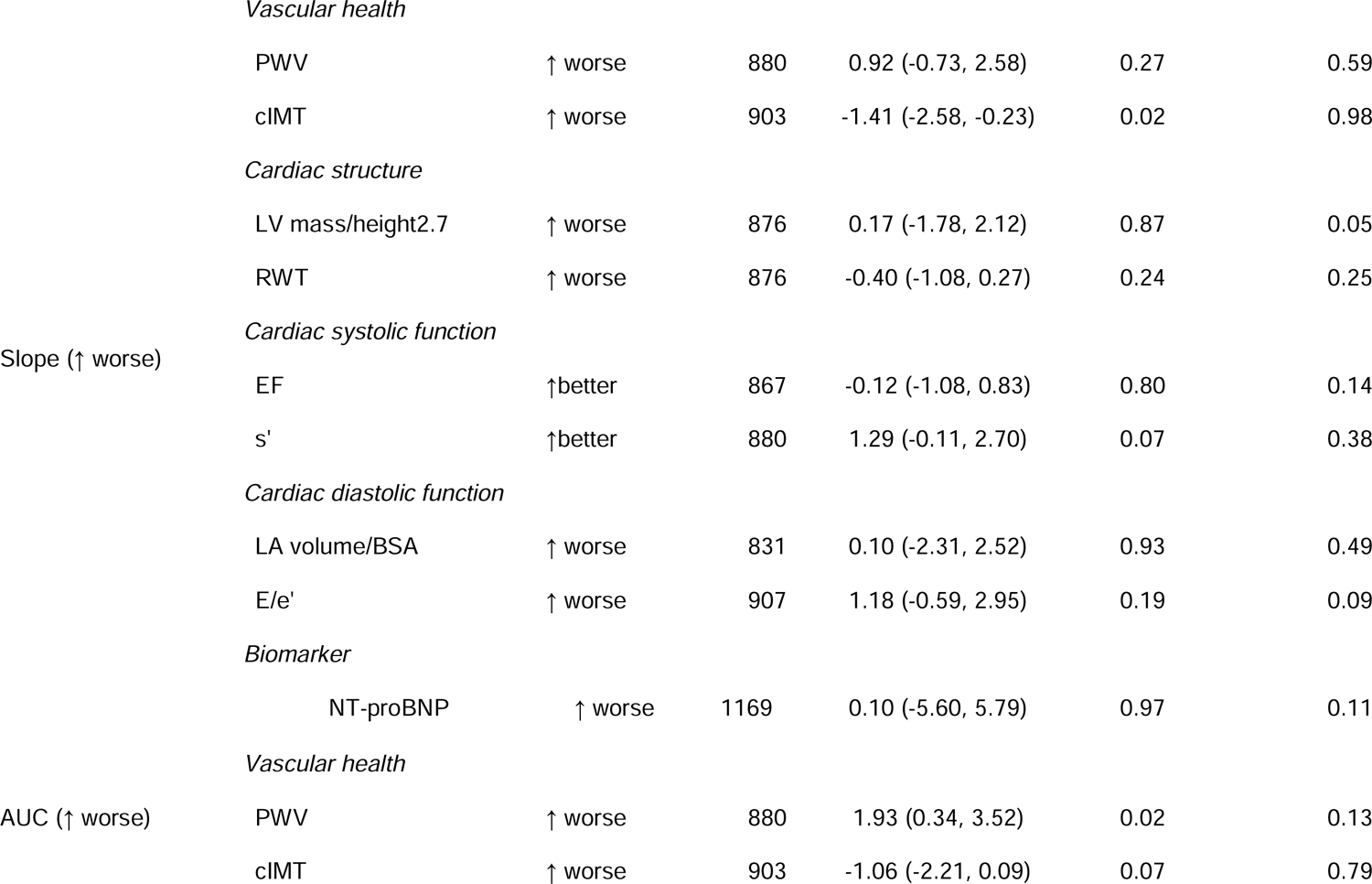
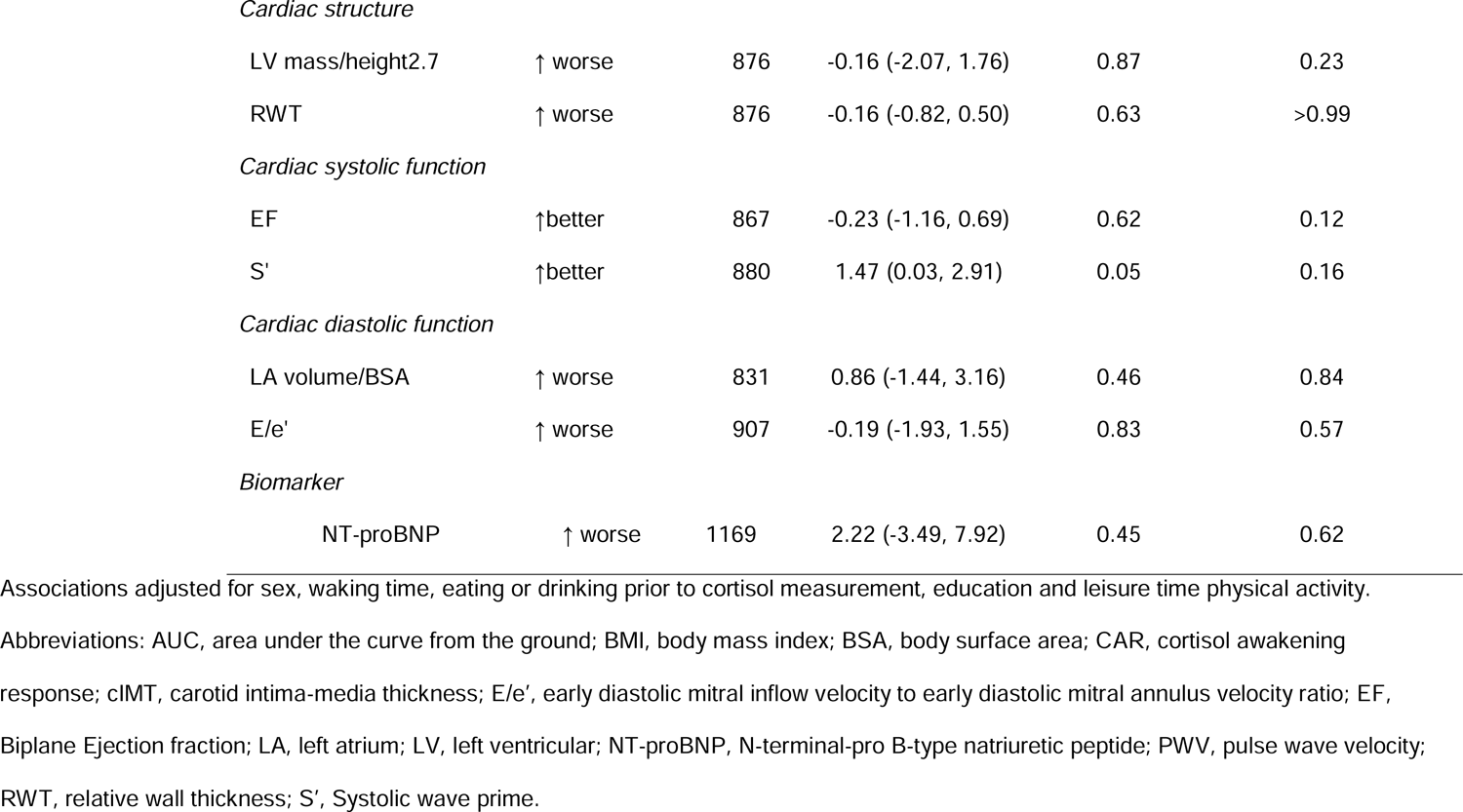
Adjusted associations of cortisol measures (per SD) and cardiovascular parameters.

**Table 3:** Association of cortisol measures (per SD) and cardiac remodelling categories.

|  | N | CAR (↑ better) |  | Slope (↑ worse) |  | AUC (↑ worse) |  |
| --- | --- | --- | --- | --- | --- | --- | --- |
|  |  | OR (95%-CI) | P value | OR (95%-CI) | P value | OR (95%-CI) | P value |
| *Remodelling categories* |  |  |  |  |  |  |  |
| normal | 380 | Ref. (1.00) |  | Ref. (1.00) |  | Ref. (1.00) |  |
| concentric hypertrophy | 143 | 1.02 (0.84, 1.23) | 0.87 | 0.97 (0.80, 1.19) | 0.79 | 0.93 (0.75, 1.14) | 0.47 |
| eccentric hypertrophy | 128 | 0.86 (0.70, 1.05) | 0.15 | 1.00 (0.81, 1.23) | 0.99 | 1.06 (0.88, 1.27) | 0.54 |
| concentric remodelling | 225 | 0.88 (0.75, 1.05) | 0.15 | 0.83 (0.70, 0.99) | 0.03 | 0.87 (0.72, 1.05) | 0.14 |

Adjusted for adjusted for sex, waking time, eating or drinking prior to cortisol measurement, education and leisure time physical activity. P for sex interactions were all >0.3.
Abbreviations: AUC, area under the curve from the ground; CAR, cortisol awakening response.

In sensitivity analyses excluding potential outliers, there was an additional inverse association of AUC and cIMT (−1.2%, 95%-CI −2.3, −0.1) and AUC was not associated with s’, suggesting these associations were driven by outliers. In sensitivity analyses excluding people with prevalent myocardial infarction and angina, there was an additional inverse association of CAR and PWV (−1.90%, 95%-CI −3.75, −0.05), and all other associations from the main analyses were similar (Supplementary Table S4).

Linked data were available for 1069 participants. Over a mean 12 years of follow-up there were 104 total deaths, 23 cardiovascular and 76 non-cardiovascular deaths. There were no clear associations between CAR, cortisol slope, and AUC with all-cause, cardiovascular and non-cardiovascular mortality in models adjusted for sex, any eating or drinking, waking time, education, and physical activity (Figure 1). However, we note that the precision of these estimates was limited due to few disease events in the sample. Associations of AUC with outcomes showed an inverse direction. Results did not substantially differ when fewer covariates were included and after additional adjustment for BMI and Framingham risk score, and showed no evidence for non-linearity (*P* value >0.09) and sex differences (*P* value >0.2). In sensitivity analyses, the overall results for the association of cortisol measures and mortality did not differ (Supplementary Table S5).

**Figure 1:**
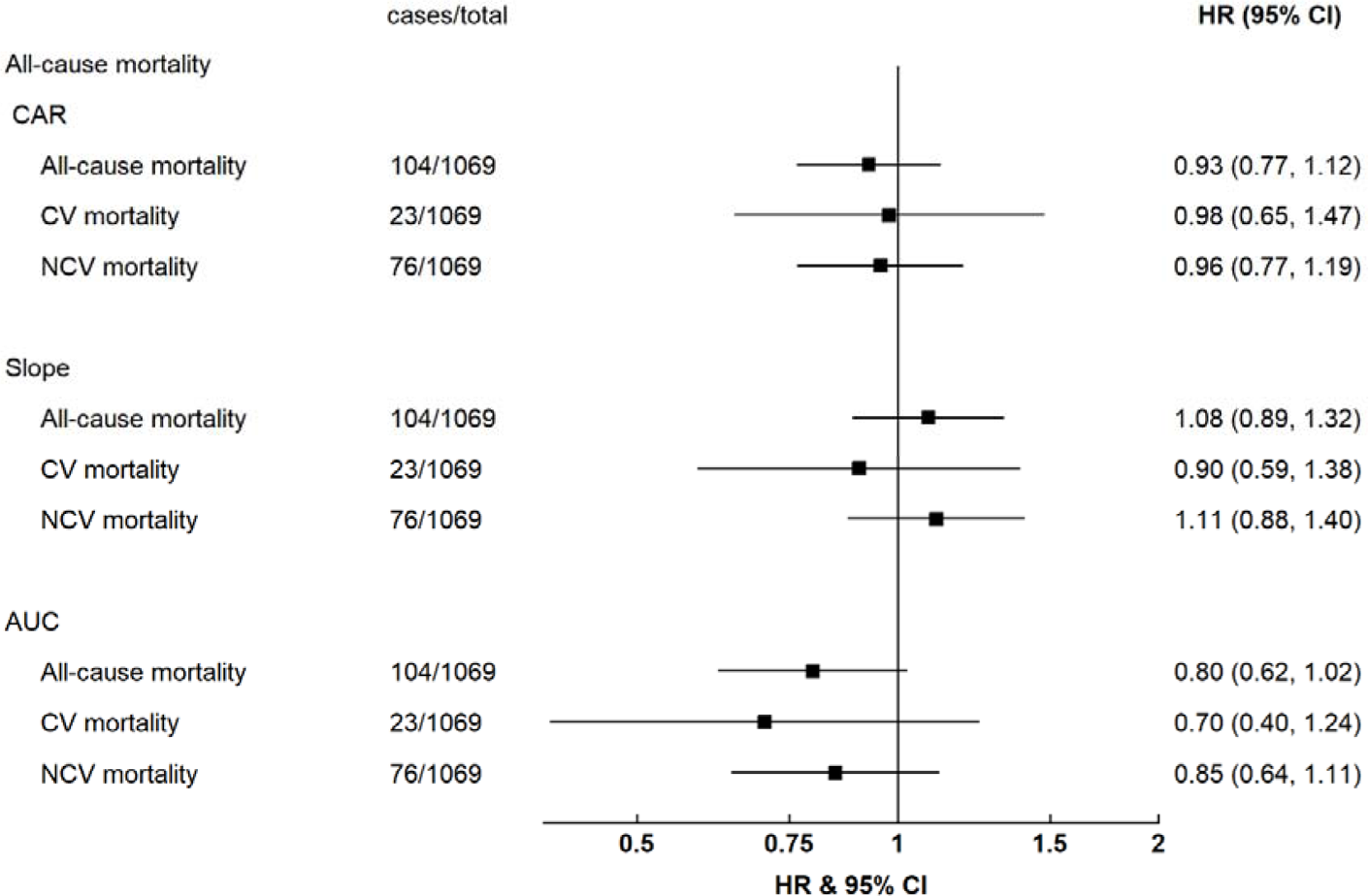
Risk of all-cause, cardiovascular (CV), and non-cardiovascular (NCV) mortality per SD higher cortisol-awakening response (CAR), cortisol slope and area under the curve (AUC) at age 60 to 64 years

## 5. Discussion

In this study of older British adults, there was little evidence that variation in diurnal cortisol was associated with subclinical cardiovascular disease or incidence cardiovascular mortality. The observed positive associations of average daily cortisol (AUC) with arterial stiffness, and lower cIMT, and shallow cortisol slope with lower cIMT and lower likelihood of concentric remodelling, were very weak.

Our findings add to current knowledge on the relationship between cortisol and atherosclerosis or arterial stiffness (arteriosclerosis). Two studies of participants around the ages of 40 (sample sizes 718 and 488) had previously found an association between a flatter cortisol slope and higher cIMT and risk of coronary artery calcification (CAC). ^11,15^ A study in 1866 older participants (mean 75 years) found AUC to be associated with a higher plaque score.^10^ However, a study of 610 participants in the Multi-Ethnic Study of Atherosclerosis of similar age to our study found weak and directionally inconsistent associations between cortisol and two subclinical measure of atherosclerosis (CAC and brachial ankle index),^12^ and a study of 367 women in the Coronary Artery Risk Development in Young Adults study found no association between AUC or slope and CAC or cIMT.^13^ Apart from Dekker *et al.*,^10^ no other study found an association of CAR and AUC with measures of subclinical atherosclerosis.^11–13,15^ A longitundinal study of 3281 participants of ∼65 years of age found no associations between salivary cortisol and PWV but observed that a more shallow salivary cortisol slope at baseline was associated with a greater change in PWV over the subsequent 5 years in women but not in men.^14^ Hajat *et al.* hypothesised that cortisol might be more relevant in the development of atherosclerosis at younger ages and of lesser importance in older age.^12^ However, this view is not be supported by a Mendelian randomisation study which used genetic variants to index long-term exposure to circulating cortisol, and found no evidence for effects of cortisol exposure on cardiometabolic diseases.^28^ We also note that previous studies on cortisol and CVD measures to have reported associations in line with the hypothesis of the role of HPA dysregulation on CVD outcomes were often limited to few subclinical measures or showed associations with only few measures of cortisol pattern and some results being limited to subgroups.^6,8,10–15^ Another consideration is that the evidence might be affected by publication bias with unexpected or null findings not yielding peer-reviewed publications.

To the best of our knowledge, this is the first study to report the association of salivary cortisol and heart function and structure. We found little or no convincing evidence of relationships: a weak positive association with s’ was driven by outliers, and an association of a more shallow slope with lower odds of concentric remodelling was unexpected and inconsistent with associations with PWV. Concentric remodelling can occur in response to chronic pressure, overload or MI.^29^ However, we cannot rule out that this was a chance finding due to multiple testing and potentially exacerbated by reduced power due to multinomial analyses requiring large sample sizes.

We found no evidence of an association of diurnal cortisol measures with CV or non-CV mortality. However, estimates were imprecise and we cannot exclude the possibility of clinically important associations. A recent study of 1090 participants of a representative German sample (mean age 60) including 87 deaths (of which 31 were cardiovascular) had found no suggestion of an association with AUC.^8^ Across all cortisol measures, previous evidence is mixed. The aforementioned study found an inverse association of CAR, and peak-to-bedtime ratio, reflecting a steeper slope and lower risk of CVD deaths and a study of 4047 participants of the Whitehall II study, with 139 total and 32 CV deaths, showed a more shallow salivary cortisol slope to be associated with greater CVD mortality risk ^6^. However, a study of older adults (mean 76 years) including 85 fatal CHD cases found no association with cortisol slope.^9^

Strengths of the study were the long follow-up (12 years) for analyses of mortality and the use of a large array of detailed subclinical cardiovascular measures. The availability of several salivary cortisol measures across the day allowed us to investigate diurnal cortisol rhythm including CAR, diurnal cortisol slope as well as the average level across the day.^2^ e also emphasise several limitations. First, we cannot exclude the possibility that associations found might have occurred by chance (false positives), especially considering multiple testing in relation to the large number of exposure-outcome analyses. Conversely, precision for detecting associations was clearly limited in some circumstances – notably the analyses of incident mortality risk (raising the possibility of false negative findings). Cortisol was only measured on four occasions over 24 hours which could introduce measurement error that would bias results towards the null, especially for models involving AUC. Cortisol measurements have large day-to-day variations,^30^ and may be influenced by emotional experiences on the day, or preceding day of measurement.^31^ A study comparing the estimation of cortisol slope, CAR and AUC using fewer than an hourly measurement protocol suggested that slope and CAR can be estimated from as little as two measurements without loss of information, but that AUC estimations of fewer than hourly measurements were less well correlated.^32^ Another consideration is that these analyses are observational and results might have been affected by residual confounding and reverse causation. The findings might also have been affected by selection biases: it is possible that participants with more HPA dysregulation were more likely to be missing from the sample, and there might also be competing risks affecting mortality associations. Finally, generalisability might be limited by the study sample which is exclusively of European ethnicity and shows a healthy cohort effect with more advantaged participants having remained in follow-up over the decades of the study, and given that these participants are all of a single age, there may also be secular differences in these associations in other generations.

### Future directions

In conclusion, this study does not provide convincing evidence for a clinically important link between HPA dysregulation and subclinical cardiovascular disease or cardiovascular mortality. However, future studies using a broad array of cardiovascular measures, as well as a larger burden of incident disease could address the current limitations of the evidence base to further characterise the relative importance of the cortisol pathway for CVD.

## Data Availability

Data used in this publication are available to bona fide researchers upon request to the NSHD Data Sharing Committee via a standard application procedure. Further details can be found at http://www.nshd.mrc.ac.uk/data

## Acknowledgements

We would like to thank the NSHD participants and the numerous team members involved in the study including interviewers, technicians, researchers, administrators, managers, health professionals and volunteers. We are additionally grateful to our funders for their financial input and support in making this research happen.

