## Supplemental tables for "Associations of salivary cortisol with cardiovascular parameters and mortality in the MRC National Survey of Health and Development (NSHD)"

### Supplementary material

Table S1: Sample characteristics by inclusion into analytical sample vs. participants from the wider NSHD not eligible for analyses

| **Characteristic** | **Included sample** | | **Excluded sample** | | **p-value** |
| --- | --- | --- | --- | --- | --- |
|  | N | n (%)/*mean (SD)* | N | n (%)/*mean (SD)* |  |
| PWV distance/time (m/s) | 880 | *8.35 (2.68)* | 409 | *8.59 (2.90)* | 0.14 |
| cIMT (mm) | 903 | *0.68 (0.12)* | 633 | *0.69 (0.13)* | 0.45 |
| LV mass indexed for height^2.7^ (g/m^2.7^) | 876 | *44.32 (13.80)* | 604 | *45.07 (15.98)* | 0.33 |
| RWT | 876 | *0.42 (0.10)* | 608 | *0.43 (0.14)* | 0.02 |
| EF (%) | 867 | *64.29 (7.95)* | 587 | *64.22 (7.74)* | 0.86 |
| S' (cm/s) | 880 | *7.79 (1.61)* | 632 | *7.77 (1.69)* | 0.78 |
| LA volume/BSA (ml/m^2^) | 831 | *23.47 (8.17)* | 593 | *23.77 (7.80)* | 0.50 |
| E/e' (%) | 907 | *7.92 (2.16)* | 659 | *7.90 (2.14)* | 0.86 |
| NT-proBNP (pg/ml) | 1,169 | *91.82 (168.84)* | 885 | *92.77 (166.67)* | 0.9 |
| Remodelling | 876 |  | 604 |  | 0.54 |
| normal |  | 380 (43.38) |  | 249 (41.23) |  |
| concentric hypertrophy |  | 143 (16.32) |  | 105 (17.38) |  |
| eccentric hypertrophy |  | 128 (14.61) |  | 79 (13.08) |  |
| concentric remodelling |  | 225 (25.68) |  | 171 (28.31) |  |
| Any eating/drinking prior to cortisol measurement |  |  |  |  |  |
| no | 1,263 | 1,074 (85.04) | 469 | 302 (64.39) | <0.001 |
| yes |  | 189 (14.96) |  | 167 (35.61) |  |
| Waking time |  |  |  |  |  |
| before 6am | 1,263 | 144 (11.40) | 523 | 58 (11.09) | <0.01 |
| 6-6.30am |  | 172 (13.62) |  | 59 (11.28) |  |
| 6.30-7am |  | 243 (19.24) |  | 76 (14.53) |  |
| 7-7.30am |  | 257 (20.35) |  | 99 (18.93) |  |
| 7.30-8am |  | 197 (15.60) |  | 86 (16.44) |  |
| 8-8.30am |  | 132 (10.45) |  | 62 (11.85) |  |
| after 8.30am |  | 118 (9.34) |  | 83 (15.87) |  |
| Sex | 1,263 |  | 4,097 |  |  |
| women |  | 590 (46.71) |  | 2,224 (54.28) | <0.001 |
| men |  | 673 (53.29) |  | 1,873 (45.72) |  |
| Highest educational attainment | 1,263 |  | 3,356 |  |  |
| <O level/equivalent |  | 419 (33.17) |  | 1,718 (51.19) | <0.001 |
| O level/equivalent |  | 248 (19.64) |  | 587 (17.49) |  |
| ≥A level/equivalent |  | 596 (47.19) |  | 1,051 (31.32) |  |
| Leisure time physical activity | 1,263 |  | 925 |  |  |
| none |  | 784 (62.07) |  | 616 (66.59) | 0.06 |
| 1-4times/week |  | 188 (14.89) |  | 111 (12.00) |  |
| 5+times/week |  | 291 (23.04) |  | 198 (21.41) |  |
| Framingham Risk Score | 1,049 | *6.83 (2.47)* | 739 | *7.33 (2.68)* | <0.001 |
| BMI kg/m^2^ | 1,260 | *27.88 (4.95)* | 959 | *27.99 (4.80)* | 0.58 |

Abbreviations: BMI, body mass index; BSA, body surface area; cIMT, carotid intima-media thickness; E/e’, early diastolic mitral inflow velocity to early diastolic mitral annulus velocity ratio; EF, Biplane Ejection fraction; LA, left atrium; LV, left ventricular; NT-proBNP, N-terminal-pro B-type natriuretic peptide; PWV, pulse wave velocity; RWT, relative wall thickness; S’, Systolic wave prime.

Table S2: Associations of cortisol measures (per SD) and cardiovascular parameters by adjusted model

| Cortisol measure | Cardiovascular parameter |  | Model 1 | | | Model 2 | | | | Model 3 | | | |  | | Model 4 | | |
| --- | --- | --- | --- | --- | --- | --- | --- | --- | --- | --- | --- | --- | --- | --- | --- | --- | --- | --- |
|  |  | **N** | % difference (95%-CI) / *OR (95%-CI)* | P value | % difference (95%-CI) / *OR (95%-CI)* | | | P value | % difference (95%-CI) / *OR (95%-CI)* | | P value | | **N** | | % difference (95%-CI) / *OR (95%-CI)* | | | P value |
| CAR | *Vascular health* |  |  |  |  | | |  |  | |  | |  | |  | | |  |
|  | PWV | 880 | -1.56 (-3.16, 0.03) | 0.05 | -1.56 (-3.15, 0.04) | | | 0.06 | -1.57 (-3.16, 0.02) | | 0.05 | | 771 | | -1.51 (-3.23, 0.20) | | | 0.08 |
|  | cIMT | 903 | -0.12 (-1.29, 1.05) | 0.83 | -0.09 (-1.26, 1.08) | | | 0.88 | -0.14 (-1.30, 1.02) | | 0.81 | | 787 | | -0.34 (-1.57, 0.89) | | | 0.59 |
|  | *Cardiac structure* |  |  |  |  | | |  |  | |  | |  | |  | | |  |
|  | LV mass/height2.7 | 876 | -0.21 (-2.09, 1.67) | 0.82 | -0.16 (-2.04, 1.72) | | | 0.87 | 0.21 (-1.44, 1.86) | | 0.80 | | 759 | | -0.66 (-2.62, 1.31) | | | 0.51 |
|  | RWT | 876 | 0.10 (-0.55, 0.75) | 0.76 | 0.08 (-0.57, 0.73) | | | 0.80 | 0.10 (-0.54, 0.75) | | 0.75 | | 759 | | 0.18 (-0.54, 0.90) | | | 0.63 |
|  | *Remodelling categories* |  |  |  |  | | |  |  | |  | |  | |  | | |  |
|  | normal | 380 | *Ref. (1.00)* |  | *Ref. (1.00)* | | |  | *Ref. (1.00)* | |  | | 380 | | | | *Ref. (1.00)* |  |
|  | concentric hypertrophy | 143 | *1.02 (0.84, 1.23)* | 0.87 | *1.02 (0.84, 1.23)* | | | 0.87 | *1.03 (0.85, 1.26)* | | 0.73 | | 143 | | *0.98 (0.79, 1.22)* | | | 0.88 |
|  | eccentric hypertrophy | 128 | *0.85 (0.69, 1.04)* | 0.11 | *0.86 (0.70, 1.05)* | | | 0.15 | *0.87 (0.71, 1.07)* | | 0.19 | | 128 | | *0.82 (0.66, 1.03)* | | | 0.09 |
|  | concentric remodelling | 225 | *0.89 (0.75, 1.05)* | 0.17 | *0.88 (0.75, 1.05)* | | | 0.15 | *0.89 (0.75, 1.05)* | | 0.16 | | 225 | | *0.90 (0.75, 1.08)* | | | 0.26 |
|  | *Cardiac systolic function* |  |  |  | | |  |  |  | |  | |  | |  | | |  |
|  | EF | 867 | -0.26 (-1.19, 0.66) | 0.58 | -0.28 (-1.20, 0.65) | | | 0.56 | -0.29 (-1.22, 0.64) | | 0.54 | | 747 | | -0.50 (-1.54, 0.54) | | | 0.34 |
|  | s' | 880 | 0.02 (-1.33, 1.37) | 0.97 | -0.04 (-1.39, 1.32) | | | 0.96 | -0.04 (-1.39, 1.32) | | 0.96 | | 768 | | -0.03 (-1.50, 1.44) | | | 0.97 |
|  | *Cardiac diastolic function* |  |  |  | | |  |  |  | |  | |  | |  | | |  |
|  | LA volume/BSA | 831 | 0.44 (-1.84, 2.71) | 0.71 | 0.51 (-1.77, 2.78) | | | 0.66 | 0.61 (-1.63, 2.86) | | 0.59 | | 709 | | 0.28 (-2.19, 2.75) | | | 0.82 |
|  | E/e' | 907 | 0.07 (-1.62, 1.77) | 0.93 | 0.15 (-1.54, 1.85) | | | 0.86 | 0.19 (-1.46, 1.85) | | 0.82 | | 786 | | -0.17 (-2.00, 1.66) | | | 0.85 |
|  | *Biomarker* |  |  |  |  | | |  |  | |  | |  | |  | | |  |
|  | NT-proBNP | 1169 | -1.29 (-6.80, 4.23) | 0.65 | -1.25 (-6.77, 4.28) | | | 0.66 | -1.19 (-6.74, 4.35) | | 0.67 | | 1039 | | -1.44 (-7.37, 4.48) | | | 0.63 |
| Slope | *Vascular health* |  |  |  |  | | |  |  | |  | |  | |  | | |  |
|  | PWV | 880 | 0.92 (-0.73, 2.58) | 0.27 | 0.92 (-0.73, 2.58) | | | 0.27 | 0.84 (-0.81, 2.50) | | 0.32 | | 771 | | 0.81 (-0.98, 2.60) | | | 0.37 |
|  | cIMT | 903 | -1.39 (-2.56, -0.22) | 0.02 | -1.41 (-2.58, -0.23) | | | 0.02 | -1.47 (-2.63, -0.30) | | 0.01 | | 787 | | -1.37 (-2.61, -0.12) | | | 0.03 |
|  | *Cardiac structure* |  |  |  |  | | |  |  | |  | |  | |  | | |  |
|  | LV mass/height^2.7^ | 876 | 0.15 (-1.81, 2.10) | 0.88 | 0.17 (-1.78, 2.12) | | | 0.86 | -0.22 (-1.93, 1.49) | | 0.80 | | 759 | | 0.12 (-1.94, 2.17) | | | 0.91 |
|  | RWT | 876 | -0.40 (-1.07, 0.27) | 0.25 | -0.40 (-1.08, 0.27) | | | 0.24 | -0.43 (-1.10, 0.24) | | 0.21 | | 759 | | -0.55 (-1.31, 0.20) | | | 0.15 |
|  | *Remodelling categories* |  |  |  |  | | |  |  | |  | |  | |  | | |  |
|  | normal | 380 | *Ref. (1.00)* |  | *Ref. (1.00)* | | |  | *Ref. (1.00)* | | |  | 380 | | *Ref. (1.00)* | | |  |
|  | concentric hypertrophy | 143 | *0.96 (0.79, 1.17)* | 0.71 | *0.97 (0.80, 1.19)* | | | 0.80 | *0.96 (0.78, 1.18)* | | 0.68 | | 143 | | *0.94 (0.75, 1.18)* | | | 0.62 |
|  | eccentric hypertrophy | 128 | *0.99 (0.81, 1.22)* | 0.93 | *1.00 (0.81, 1.23)* | | | 0.98 | *0.99 (0.80, 1.22)* | | 0.91 | | 128 | | *0.92 (0.73, 1.15)* | | | 0.46 |
|  | concentric remodelling | 225 | *0.83 (0.70, 0.99)* | 0.04 | *0.83 (0.70, 0.99)* | | | 0.03 | *0.83 (0.69, 0.99)* | | 0.03 | | 225 | | *0.81 (0.67, 0.98)* | | | 0.03 |
|  | *Cardiac systolic function* |  |  |  | | |  |  |  | |  | |  | |  | | |  |
|  | EF | 867 | -0.19 (-1.14, 0.77) | 0.70 | -0.12 (-1.08, 0.83) | | | 0.80 | -0.09 (-1.04, 0.87) | | 0.86 | | 747 | | -0.42 (-1.49, 0.65) | | | 0.44 |
|  | s' | 880 | 1.34 (-0.07, 2.74) | 0.06 | 1.29 (-0.11, 2.70) | | | 0.07 | 1.29 (-0.11, 2.70) | | 0.07 | | 768 | | 1.47 (-0.06, 2.99) | | | 0.06 |
|  | *Cardiac diastolic function* |  |  |  | | |  |  |  | |  | |  | |  | | |  |
|  | LA volume/BSA | 831 | -0.04 (-2.45, 2.36) | 0.97 | 0.10 (-2.31, 2.52) | | | 0.93 | -0.16 (-2.55, 2.22) | | 0.89 | | 709 | | 0.80 (-1.85, 3.45) | | | 0.55 |
|  | E/e' | 907 | 1.13 (-0.65, 2.90) | 0.21 | 1.18 (-0.59, 2.95) | | | 0.19 | 1.00 (-0.74, 2.73) | | 0.26 | | 786 | | 0.92 (-1.01, 2.85) | | | 0.35 |
|  | *Biomarker* |  |  |  |  | | |  |  | |  | |  | |  | | |  |
|  | NT-proBNP | 1169 | 0.34 (-5.35, 6.02) | 0.91 | 0.10 (-5.60, 5.79) | | | 0.97 | 0.01 (-5.71, 5.73) | | 1.00 | | 1039 | | 0.88 (-5.34, 7.10) | | | 0.78 |
| AUC | *Vascular health* |  |  |  |  | | |  |  | |  | |  | |  | | |  |
|  | PWV | 880 | 1.79 (0.20, 3.38) | 0.03 | 1.93 (0.34, 3.52) | | | 0.02 | 1.86 (0.28, 3.44) | | 0.02 | | 771 | | 2.74 (0.75, 4.73) | | | 0.01 |
|  | cIMT | 903 | -1.09 (-2.24, 0.06) | 0.06 | -1.06 (-2.21, 0.09) | | | 0.07 | -1.11 (-2.25, 0.04) | | 0.06 | | 787 | | -1.95 (-3.36, -0.54) | | | 0.01 |
|  | *Cardiac structure* |  |  |  |  | | |  |  | |  | |  | |  | | |  |
|  | LV mass/height2.7 | 876 | -0.35 (-2.27, 1.57) | 0.72 | -0.16 (-2.07, 1.76) | | | 0.87 | -0.16 (-1.84, 1.52) | | 0.85 | | 759 | | -0.31 (-2.69, 2.08) | | | 0.80 |
|  | RWT | 876 | -0.18 (-0.83, 0.48) | 0.60 | -0.16 (-0.82, 0.50) | | | 0.63 | -0.16 (-0.82, 0.50) | | 0.63 | | 759 | | -0.27 (-1.15, 0.60) | | | 0.54 |
|  | *Remodelling categories* |  |  |  |  | | |  |  | |  | |  | |  | | |  |
|  | normal | 380 | *Ref. (1.00)* |  | *Ref. (1.00)* | | |  | *Ref. (1.00)* | |  | | 380 | | *Ref. (1.00)* | | |  |
|  | concentric hypertrophy | 143 | *0.91 (0.74, 1.12)* | 0.38 | *0.93 (0.75, 1.14)* | | | 0.47 | *0.93 (0.75, 1.15)* | | 0.49 | | 143 | | *0.89 (0.68, 1.15)* | | | 0.37 |
|  | eccentric hypertrophy | 128 | *1.04 (0.87, 1.25)* | 0.68 | *1.06 (0.88, 1.27)* | | | 0.53 | *1.06 (0.88, 1.28)* | | 0.54 | | 128 | | *0.88 (0.67, 1.15)* | | | 0.35 |
|  | concentric remodelling | 225 | *0.87 (0.73, 1.05)* | 0.15 | *0.87 (0.72, 1.05)* | | | 0.14 | *0.87 (0.72, 1.05)* | | 0.14 | | 225 | | *0.78 (0.62, 0.98)* | | | 0.03 |
|  | *Cardiac systolic function* |  |  |  | | |  |  |  | |  | |  | |  | | |  |
|  | EF | 867 | -0.29 (-1.22, 0.63) | 0.53 | -0.23 (-1.16, 0.69) | | | 0.62 | -0.22 (-1.14, 0.71) | | 0.65 | | 747 | | -0.57 (-1.79, 0.65) | | | 0.36 |
|  | s' | 880 | 1.53 (0.09, 2.97) | 0.04 | 1.47 (0.03, 2.91) | | | 0.05 | 1.47 (0.03, 2.92) | | 0.05 | | 768 | | 1.65 (-0.08, 3.39) | | | 0.06 |
|  | *Cardiac diastolic function* |  |  |  | | |  |  |  | |  | |  | |  | | |  |
|  | LA volume/BSA | 831 | 0.65 (-1.65, 2.95) | 0.58 | 0.86 (-1.44, 3.16) | | | 0.46 | 0.70 (-1.57, 2.97) | | 0.55 | | 709 | | 1.41 (-1.56, 4.38) | | | 0.35 |
|  | E/e' | 907 | -0.38 (-2.12, 1.37) | 0.67 | -0.19 (-1.93, 1.55) | | | 0.83 | -0.21 (-1.92, 1.49) | | 0.81 | | 786 | | 0.37 (-1.85, 2.60) | | | 0.74 |
|  | *Biomarker* |  |  |  |  | | |  |  | |  | |  | |  | | |  |
|  | NT-proBNP | 1169 | 2.03 (-3.67, 7.73) | 0.49 | 2.22 (-3.49, 7.92) | | | 0.45 | 2.20 (-3.51, 7.92) | | 0.45 | | 1039 | | 2.89 (-4.29, 10.06) | | | 0.43 |

Model 1: adjusted for sex, waking time, eating or drinking prior to cortisol measurement; Model 2: Model 1 additionally adjusted for education and leisure time physical activity; Model 3: Model 2 additionally adjusted for BMI; Model 4: Model 2 additionally adjusted for Framingham risk score.

Abbreviations: AUC, area under the curve from the ground; BMI, body mass index; BSA, body surface area; CAR, cortisol awakening response; cIMT, carotid intima-media thickness; E/e’, early diastolic mitral inflow velocity to early diastolic mitral annulus velocity ratio; EF, Biplane Ejection fraction; LA, left atrium; LV, left ventricular; NT-proBNP, N-terminal-pro B-type natriuretic peptide; PWV, pulse wave velocity; RWT, relative wall thickness; S’, Systolic wave prime.

Table S 3: Adjusted associations of cortisol measures (per SD) and cardiovascular parameters after exclusion of outliers (Sensitivity I) and exclusion of participants with myocardial infarction and angina (Sensitivity II)

| Cortisol measure | Cardiovascular parameter | Sensitivity I | | | | Sensitivity II | | |
| --- | --- | --- | --- | --- | --- | --- | --- | --- |
|  |  | N | % difference (95%-CI) /*OR (95%-CI)* | P value | N | | % difference (95%-CI) /*OR (95%-CI)* | P value |
| CAR | Vascular health |  |  |  |  | |  |  |
|  | PWV | 849 | -0.26 (-1.41, 0.90) | 0.66 | 739 | | -1.90 (-3.75, -0.05) | 0.04 |
|  | cIMT | 889 | -0.03 (-1.17, 1.10) | 0.95 | 754 | | -0.80 (-2.11, 0.50) | 0.23 |
|  | Cardiac structure |  |  |  |  | |  |  |
|  | LV mass/height^2.7^ | 857 | -0.36 (-2.18, 1.45) | 0.69 | 732 | | -1.22 (-3.35, 0.91) | 0.26 |
|  | RWT | 857 | 0.35 (-0.18, 0.89) | 0.19 | 732 | | 0.08 (-0.69, 0.85) | 0.84 |
|  | Remodelling categories |  |  |  |  | |  |  |
|  | normal | 380 | *Ref. (1.00)* |  | 380 | | *Ref. (1.00)* |  |
|  | concentric hypertrophy | 143 | *1.07 (0.89, 1.30)* | 0.47 | 143 | | *0.91 (0.72, 1.14)* | 0.40 |
|  | eccentric hypertrophy | 128 | *0.81 (0.66, 1.00)* | 0.05 | 128 | | *0.77 (0.61, 0.98)* | 0.04 |
|  | concentric remodelling | 225 | *0.97 (0.82, 1.14)* | 0.68 | 225 | | *0.92 (0.76, 1.11)* | 0.40 |
|  | Cardiac systolic function |  |  |  |  | |  |  |
|  | EF | 844 | 0.07 (-0.70, 0.84) | 0.86 | 723 | | -0.32 (-1.35, 0.72) | 0.55 |
|  | S' | 861 | 0.38 (-0.93, 1.69) | 0.57 | 739 | | 0.25 (-1.28, 1.77) | 0.75 |
|  | Cardiac diastolic function |  |  |  |  | |  |  |
|  | LA volume/BSA | 817 | 0.53 (-1.76, 2.82) | 0.65 | 689 | | 0.15 (-2.49, 2.79) | 0.91 |
|  | E/e' | 889 | 0.17 (-1.52, 1.86) | 0.84 | 758 | | -0.11 (-2.06, 1.83) | 0.91 |
|  | Biomarker |  |  |  |  | |  |  |
|  | NT-proBNP | 1145 | 1.55 (-3.78, 6.88) | 0.57 | 985 | | -2.11 (-8.28, 4.06) | 0.50 |
| Slope | Vascular health |  |  |  |  | |  |  |
|  | PWV | 851 | 0.81 (-0.35, 1.97) | 0.17 | 739 | | 1.17 (-0.71, 3.06) | 0.22 |
|  | cIMT | 890 | -1.35 (-2.46, -0.24) | 0.02 | 754 | | -1.38 (-2.68, -0.08) | 0.04 |
|  | Cardiac structure |  |  |  |  | |  |  |
|  | LV mass/height^2.7^ | 858 | -0.06 (-1.90, 1.78) | 0.95 | 732 | | -0.50 (-2.62, 1.63) | 0.65 |
|  | RWT | 858 | -0.38 (-0.91, 0.16) | 0.17 | 732 | | -0.66 (-1.42, 0.11) | 0.09 |
|  | Remodelling categories |  |  |  |  | |  |  |
|  | normal | 380 | *Ref. (1.00)* |  | 380 | | *Ref. (1.00)* |  |
|  | concentric hypertrophy | 143 | *0.98 (0.80, 1.18)* | 0.81 | 143 | | *0.95 (0.76, 1.20)* | 0.69 |
|  | eccentric hypertrophy | 128 | *0.86 (0.70, 1.06)* | 0.16 | 128 | | *0.90 (0.71, 1.14)* | 0.37 |
|  | concentric remodelling | 225 | *0.78 (0.66, 0.93)* | 0.005 | 225 | | *0.79 (0.65, 0.96)* | 0.02 |
|  | Cardiac systolic function |  |  |  |  | |  |  |
|  | EF | 844 | -0.18 (-0.95, 0.59) | 0.65 | 723 | | -0.29 (-1.31, 0.73) | 0.58 |
|  | S' | 863 | 0.89 (-0.40, 2.17) | 0.18 | 739 | | 1.81 (0.32, 3.30) | 0.02 |
|  | Cardiac diastolic function |  |  |  |  | |  |  |
|  | LA volume/BSA | 818 | 0.08 (-2.29, 2.44) | 0.95 | 689 | | 0.19 (-2.51, 2.88) | 0.89 |
|  | E/e' | 892 | 1.12 (-0.57, 2.82) | 0.19 | 758 | | 0.73 (-1.21, 2.67) | 0.46 |
|  | Biomarker |  |  |  |  | |  |  |
|  | NT-proBNP | 1145 | 1.86 (-3.49, 7.20) | 0.50 | 985 | | 0.78 (-5.36, 6.93) | 0.80 |
| AUC | Vascular health |  |  |  |  | |  |  |
|  | PWV | 852 | 1.56 (0.42, 2.69) | 0.007 | 739 | | 2.66 (0.80, 4.52) | 0.005 |
|  | cIMT | 893 | -1.20 (-2.28, -0.11) | 0.03 | 754 | | -1.15 (-2.47, 0.17) | 0.09 |
|  | Cardiac structure |  |  |  |  | |  |  |
|  | LV mass/height^2.7^ | 861 | 0.18 (-1.65, 2.01) | 0.85 | 732 | | -0.42 (-2.59, 1.75) | 0.70 |
|  | RWT | 861 | -0.40 (-0.93, 0.13) | 0.14 | 732 | | -0.27 (-1.05, 0.51) | 0.50 |
|  | Remodelling categories |  |  |  |  | |  |  |
|  | normal | 380 | *Ref. (1.00)* |  | 380 | | *Ref. (1.00)* |  |
|  | concentric hypertrophy | 143 | *0.92 (0.76, 1.12)* | 0.41 | 143 | | *0.93 (0.73, 1.18)* | 0.55 |
|  | eccentric hypertrophy | 128 | *0.94 (0.77, 1.15)* | 0.55 | 128 | | *1.05 (0.85, 1.30)* | 0.66 |
|  | concentric remodelling | 225 | *0.85 (0.71, 1.00)* | 0.05 | 225 | | *0.82 (0.66, 1.02)* | 0.07 |
|  | Cardiac systolic function |  |  |  |  | |  |  |
|  | EF | 846 | -0.42 (-1.17, 0.32) | 0.27 | 723 | | -0.11 (-1.12, 0.91) | 0.83 |
|  | S' | 866 | 0.90 (-0.34, 2.14) | 0.16 | 739 | | 1.54 (0.04, 3.04) | 0.04 |
|  | Cardiac diastolic function |  |  |  |  | |  |  |
|  | LA volume/BSA | 822 | 0.97 (-1.27, 3.21) | 0.40 | 689 | | 1.12 (-1.53, 3.76) | 0.41 |
|  | E/e' | 895 | 0.15 (-1.51, 1.81) | 0.86 | 758 | | 0.21 (-1.76, 2.19) | 0.83 |
|  | Biomarker |  |  |  |  | |  |  |
|  | NT-proBNP | 1148 | 5.18 (-0.14, 10.50) | 0.06 | 985 | | 5.22 (-1.08, 11.52) | 0.10 |

Associations adjusted for sex, waking time, eating or drinking prior to cortisol measurement, education and leisure time physical activity.

Abbreviations: AUC, area under the curve from the ground; BSA, body surface area; CAR, cortisol awakening response; cIMT, carotid intima-media thickness; E/e’, early diastolic mitral inflow velocity to early diastolic mitral annulus velocity ratio; EF, Biplane Ejection fraction; LA, left atrium; LV, left ventricular; NT-proBNP, N-terminal-pro B-type natriuretic peptide; PWV, pulse wave velocity; RWT, relative wall thickness; S’, Systolic wave prime.

Table S 5: Adjusted associations of cortisol measures (per SD) and mortality after exclusion of outliers (Sensitivity I), exclusion of participants with myocardial infarction and angina (Sensitivity II) and exclusion of any deaths within the first 2 years of follow-up (Sensitivity III)

|  |  | **Sensitivity I** | | **Sensitivity II** | | **Sensitivity III** | |
| --- | --- | --- | --- | --- | --- | --- | --- |
|  | | **cases/N** | **HR (95%-CI)** | **cases/N** | **HR (95%-CI)** | **cases/N** | **HR (95%-CI)** |
| CAR |  |  | |  | |  | |
| All-cause mortality | | 103 / 1053 | 0.90 (0.74, 1.10) | 77 / 897 | 0.99 (0.79, 1.25) | 101 / 1066 | 0.93 (0.76, 1.12) |
| CV mortality | | 22 / 1053 | 0.82 (0.53, 1.28) | 15 / 897 | 1.03 (0.62, 1.71) | 22 / 1066 | 0.99 (0.65, 1.50) |
| NCV mortality | | 76 / 1053 | 0.98 (0.78, 1.23) | 62 / 897 | 0.99 (0.77, 1.27) | 74 / 1066 | 0.95 (0.77, 1.19) |
| Slope |  |  |  |  |  |  |  |
| All-cause mortality | | 104 / 1055 | 1.11 (0.92, 1.35) | 77 / 897 | 1.02 (0.81, 1.30) | 101 / 1066 | 1.12 (0.92, 1.37) |
| CV mortality | | 23 / 1055 | 0.89 (0.58, 1.36) | 15 / 897 | 1.09 (0.63, 1.87) | 22 / 1066 | 0.96 (0.62, 1.48) |
| NCV mortality | | 76 / 1055 | 1.15 (0.91, 1.44) | 62 / 897 | 1.01 (0.77, 1.32) | 74 / 1066 | 1.14 (0.90, 1.44) |
| AUC |  |  |  |  |  |  |  |
| All-cause mortality | | 104 / 1055 | 0.87 (0.71, 1.07) | 77 / 897 | 0.85 (0.65, 1.12) | 101 / 1066 | 0.81 (0.63, 1.04) |
| CV mortality | | 23 / 1055 | 0.78 (0.49, 1.22) | 15 / 897 | 0.78 (0.40, 1.51) | 22 / 1066 | 0.78 (0.45, 1.35) |
| NCV mortality | | 76 / 1055 | 0.92 (0.73, 1.17) | 62 / 897 | 0.86 (0.64, 1.17) | 74 / 1066 | 0.84 (0.64, 1.11) |

Associations adjusted for sex, waking time, eating or drinking prior to cortisol measurement, education and leisure time physical activity.

Abbreviations: AUC, area under the curve from the ground; CAR, cortisol awakening response; cIMT, carotid intima-media thickness.
